# Fine-Tuning PubMedBERT for Hierarchical Condition Category Classification from Clinical Notes

**DOI:** 10.64898/2026.04.13.26350814

**Authors:** Xiangren Wang, Noah Hammarlund, Mattia Prosperi, Yenan Zhu, Lee Revere

## Abstract

Automating Hierarchical Condition Category (HCC) assignment directly from unstructured electronic health record (EHR) notes remains an important but understudied problem in clinical informatics. We present HCC-Coder, an end-to-end NLP system that maps narrative documentation to 115 Centers for Medicare & Medicaid Services(CMS) HCC codes in a multi-label setting. On the test dataset, HCC-Coder achieves a macro-F1 of 0.779 and a micro-F1 of 0.756, with a macro-sensitivity of 0.819 and macro-specificity of 0.998. By contrast, Generative Pre-trained Transformer (GPT)-4o achieves the highest score of a macro-F1 of 0.735 and a micro-F1 of 0.708 under five-shot prompting. The fine-tuned model demonstrates consistent absolute improvements of 4%–5% in F1-scores over GPT-4o. To address severe label imbalance, we incorporate inverse-frequency weighting and per-label threshold calibration. These findings suggest that domain-adapted transformers provide more balanced and reliable performance than prompt-based large language models for hierarchical clinical coding and risk adjustment.

## Introduction

Hierarchical Condition Category (HCC) codes are a cornerstone of risk adjustment systems used by the Centers for Medicare & Medicaid Services (CMS) to ensure fair reimbursement and equitable care delivery.^1^ These codes summarize patients’ chronic conditions and disease severity, directly influencing healthcare payment models and population-level health management.^1^ However, the current HCC assignment relies heavily on manual review of clinical documentation, making the process time-consuming, inconsistent, and difficult to scale in large health systems.

Recent advances in biomedical natural language processing (NLP) have opened new opportunities to automate this process. Transformer-based language models such as BioBERT,^2^ ClinicalBERT,^3^ and PubMedBERT^4^ have demonstrated strong performance in a range of clinical text-mining tasks, including entity recognition, phenotyping, and diagnosis classification.^5^ Nevertheless, relatively limited work has focused on large-scale, multi-label, hierarchical tasks such as HCC classification, where individual clinical notes may correspond to multiple chronic conditions with highly imbalanced prevalence.

To address these challenges, we introduce HCC-Coder, a transformer-based framework that fine-tunes PubMedBERT for large-scale, multi-label classification of 115 HCC codes from unstructured electronic health record (HER) notes. We develop a semi-supervised labeling pipeline that leverages Generative Pre-trained Transformer (GPT)-based large language models guided by official CMS definitions and subsequently verified by domain experts.^6^ In addition, we incorporate inverse-frequency weighting and per-label threshold calibration to improve model calibration and interpretability. Through a comprehensive evaluation on a dataset derived from MIMIC-IV,^7^ HCC-Coder outperforms GPT-4o under both zero-shot and five-shot prompting, demonstrating the potential of domain-specific transformer models for automating healthcare risk adjustment.

## Related Work

### Automated Medical Coding

Automated medical coding seeks to map free-text clinical documentation to standardized taxonomies such as diagnosis codes, billing codes, and health behaviors.^**8**, **9**, **10**, **11**, **12**^ Early approaches relied on rule-based systems and feature engineering, including TF–IDF, bag-of-words, and keyword matching pipelines.^**13**^ While these methods provided transparency, they were limited by vocabulary sparsity and the inability to model semantic context. Subsequent neural approaches introduced convolutional and recurrent architectures that learned distributed representations of clinical notes.^**14**^ However, these models struggled with long-range dependencies and label imbalance, leading to underperformance on rare or hierarchical disease categories.

### Transformer Models for Clinical NLP

Transformers have advanced clinical NLP via contextual embeddings and transfer learning, with BioBERT,^**2**^ ClinicalBERT,^**3**^ and PubMedBERT^**4**^ improving phenotyping and note classification. Yet most work targets single-label or sentence-level tasks, not large-scale multi-label problems like risk adjustment, and the field still lacks standardized, annotated datasets for hierarchical coding. Meanwhile, general-purpose LLMs are designed primarily as generative models rather than dedicated discriminative classifiers and are not explicitly optimized for hierarchical multilabel prediction tasks such as HCC coding. As a result, their outputs may require additional prompting and calibration to achieve stable label-wise decision boundaries, particularly under severe class imbalance. These gaps motivate lightweight, domain-adapted transformers that finetune efficiently, calibrate well, and are practical to reproduce and deploy.

### Position of This Work

Building on prior transformer-based research in medical coding, we explore the use of PubMedBERT for large-scale, multi-label HCC classification from unstructured HER notes. Our focus is on addressing practical challenges inherent to hierarchical risk-adjustment coding, particularly severe label imbalance and heterogeneous prevalence across categories. By integrating inverse-frequency weighting and per-label threshold calibration within a unified evaluation framework, we provide a systematic assessment of transformer-based approaches for structured HCC coding.

## Material and Methods

### Data Description

We randomly selected approximately 36,000 de-identified critical-care HER notes from MIMIC-IV and constructed HCC ground truth via a semi-automated pipeline.^7^ ChatGPT-4o was prompted with explicit task instructions to assign all applicable CMS HCC codes to each note and was provided with the official CMS HCC coding criteria and three exemplar note–label pairs. GPT-4o achieved an exact-match of 53.7%, meaning a prediction was counted as correct only when the entire set of HCC codes exactly matched the ground truth. Given this level of performance, we conducted a full manual review of the entire dataset of 36,000 notes. The clinical researchers systematically audited and corrected every single GPT-generated label according to CMS guidelines. The dataset was then split using a multilabel-stratified strategy (70% training, 20% validation, 10% test) to preserve label prevalence across sets.

### Model Architecture

We fine-tuned the open-source PubMedBERT model, which was pre-trained on large-scale biomedical and clinical corpora. While PubMedBERT captures domain-specific language representations,^4^ supervised fine-tuning is required for downstream clinical prediction tasks such as multi-label HCC classification. For HCC prediction, we adopted a standard multi-label classification formulation. A linear classification layer with 115 output units (corresponding to the HCC codes) was applied to the final-layer [CLS] representation. Because each note may contain multiple HCC labels, output probabilities were computed independently for each label using the sigmoid function.

We optimized the model using standard binary cross-entropy loss^15^, which treats each HCC code as an independent prediction task. This formulation is widely used in multi-label text classification and is particularly appropriate in settings where multiple labels can co-occur. All PubMedBERT parameters were updated end-to-end during fine-tuning. Computational optimizations were applied to improve training efficiency without modifying the underlying model architecture. The overall model architecture is illustrated in Figure 1.

**Figure 1.**
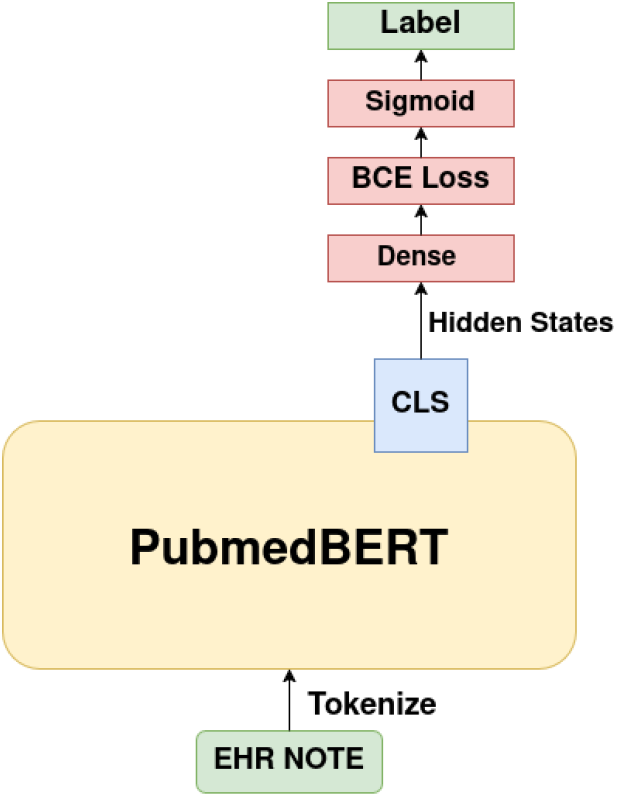
Model Architecture

### Class Imbalance Handling via Inverse-Frequency Weighting

Medical text data often exhibit substantial class imbalance, where common conditions appear frequently while many HCC categories are relatively rare. To address this imbalance, we applied inverse-frequency weighting during training, assigning greater importance to underrepresented HCC codes based on their empirical prevalence in the training set. This approach reduces bias toward frequent conditions and improves the model’s ability to detect low-prevalence categories. The resulting class weights were incorporated into the loss function during fine-tuning.

### Model Selection and Evaluation

Because our task is formulated as a multi-label classification problem rather than a single-label one, traditional accuracy is no longer an appropriate metric for model evaluation.^16^ In a single-label setting, accuracy reliably captures prediction correctness, but in a multi-label scenario, it becomes both overly strict and misleading: even a single misclassified label can render an entire sample incorrect, and the metric tends to be dominated by the majority of negative labels. This makes it insensitive to the model’s true discriminative capability across multiple co-occurring categories. Instead, we focus primarily on macro and micro F1 scores, which better reflect the balance between precision and recall in this highly imbalanced multi-label setting. In addition, we report the Area Under the ROC Curve (AUC) as a complementary, threshold-independent measure of ranking performance.^17^ Since our validation phase does not involve explicit threshold optimization for maximizing F1-score, AUC serves as a more stable and informative indicator of model quality. It inherently summarizes how well the model separates classes under all possible thresholds, effectively reflecting its overall selection and discrimination ability. During training, we saved checkpoints at every epoch and tracked validation AUC and loss. After training, we selected the checkpoint with the highest validation AUC within a stable plateau region (defined as ≤0.2 percentage-point change over three consecutive epochs) and prior to clear divergence between training and validation loss. Decision thresholds are determined exclusively on the validation set and then fixed prior to evaluation on the test set. Final F1-based evaluation results are computed on the test set using these fixed thresholds.

## Results

### Untrained PubMedBERT Baseline Performance

To establish a benchmark for comparison, we evaluated the untrained PubMedBERT model modified with our additional dense classification head but without any fine-tuning. This baseline represents the raw capability of the model with its randomly initialized classification head to perform HCC classification without task specific optimization on labeled clinical notes.

We applied the model to the validation set to assess its classification performance, performing threshold tuning on the sigmoid outputs using two distinct strategies to identify the optimal approach for subsequent test set evaluation. Specifically, thresholds were optimized on the validation set using grid search over probability values from 0.01 to 0.99 in increments of 0.05, with macro-F1 as the optimization criterion. Under the global strategy, a single threshold was applied uniformly across all labels. Under the per-label strategy, label-specific thresholds were independently optimized for each HCC code. The validation-derived thresholds were then fixed and applied to the test set without further tuning. The performance of both thresholding strategies on the validation and test sets is summarized in Tables 1A and 1B.

**Table 1A.** Baseline performance on the validation set under global and per-label threshold tuning.

| Threshold Strategy | Metric | Macro | Macro(95%CI) | Micro | Micro(95%CI) |
| --- | --- | --- | --- | --- | --- |
| Global threshold | F1 | 0.019 | 0.018-0.021 | 0.019 | 0.018-0.021 |
|  | Sensitivity | 0.957 | 0.953-0.962 | 0.953 | 0.947-0.958 |
|  | Specificity | 0.043 | 0.042-0.043 | 0.043 | 0.042-0.043 |
|  | Precision | 0.009 | 0.009-0.012 | 0.010 | 0.009-0.010 |
| Per-label thresholds | F1 | 0.029 | 0.026-0.033 | 0.028 | 0.025-0.030 |
|  | Sensitivity | 0.548 | 0.538-0.559 | 0.656 | 0.640-0.670 |
|  | Specificity | 0.549 | 0.549-0.551 | 0.551 | 0.550-0.552 |
|  | Precision | 0.022 | 0.016-0.028 | 0.014 | 0.013-0.015 |

**Table 1B.** Baseline performance on the test set using validation-selected global and per-label threshold.

| Threshold Strategy | Metric | Macro | Macro(95%CI) | Micro | Micro(95%CI) |
| --- | --- | --- | --- | --- | --- |
| Global threshold | F1 | 0.014 | 0.014-0.015 | 0.015 | 0.013-0.015 |
|  | Sensitivity | 0.959 | 0.952-0.966 | 0.953 | 0.944-0.961 |
|  | Specificity | 0.043 | 0.042-0.043 | 0.042 | 0.042-0.043 |
|  | Precision | 0.007 | 0.007-0.008 | 0.007 | 0.007-0.007 |
| Per-label thresholds | F1 | 0.017 | 0.015-0.019 | 0.018 | 0.017-0.019 |
|  | Sensitivity | 0.493 | 0.479-0.507 | 0.573 | 0.555-0.591 |
|  | Specificity | 0.549 | 0.548-0.550 | 0.550 | 0.549-0.551 |
|  | Precision | 0.010 | 0.008-0.012 | 0.009 | 0.008-0.009 |

Across both the validation (Table 1A) and test sets (Table 1B), the per-label threshold strategy consistently outperformed the global threshold approach in terms of F1 score and operating balance. On the validation set, per-label thresholding increased macro-F1 from 0.019 to 0.030 (absolute +0.011; ∼58% relative improvement). On the test set, macro-F1 similarly improved from 0.014 under the global threshold to 0.017 with per-label tuning (absolute +0.003; ∼21% relative improvement). Although the global threshold strategy achieved extremely high sensitivity (≈0.96), it was accompanied by very low specificity (≈0.04), indicating a recall-dominant operating point that over-predicts positive labels. In contrast, the per-label strategy achieved a substantially more balanced trade-off between sensitivity (≈0.49–0.55) and specificity (≈0.55) across both datasets. The consistent improvement in F1 and the more balanced operating characteristics observed on both validation and test sets indicate stable generalization without evidence of threshold overfitting. Accordingly, the per-label test set results were adopted as the baseline for subsequent comparisons with fine-tuned models.

### Training Dynamics

To better characterize the optimization process, we tracked the training and validation losses as well as the validation AUC across all epochs. Figure 2 illustrates the learning dynamics of the fine-tuned PubMedBERT model. As shown in Figure 2, the training loss decreased steadily over epochs, while the validation loss initially followed a similar downward trend before reaching its lowest range around epochs 15–20 and gradually increasing thereafter, with mild divergence from the training curve after approximately epoch 30, although validation AUC continued to improve slightly. To mitigate this, we selected checkpoints based on performance metrics rather than the final epoch. Validation AUC increased consistently, became effectively stable by epoch 25 under a ±0.2 percentage-point, 3-epoch stability criterion, and peaked at epoch 31 with a macro-AUC of 0.966. We therefore retained the checkpoint with the highest validation macro-AUC (epoch 31) as the final model for evaluation. Overall, these dynamics demonstrate stable convergence with only mild late-stage overfitting and confirm that inverse-frequency weighting (mitigating imbalance) leads to more balanced and efficient learning across the 115 HCC codes.

**Figure 2.**
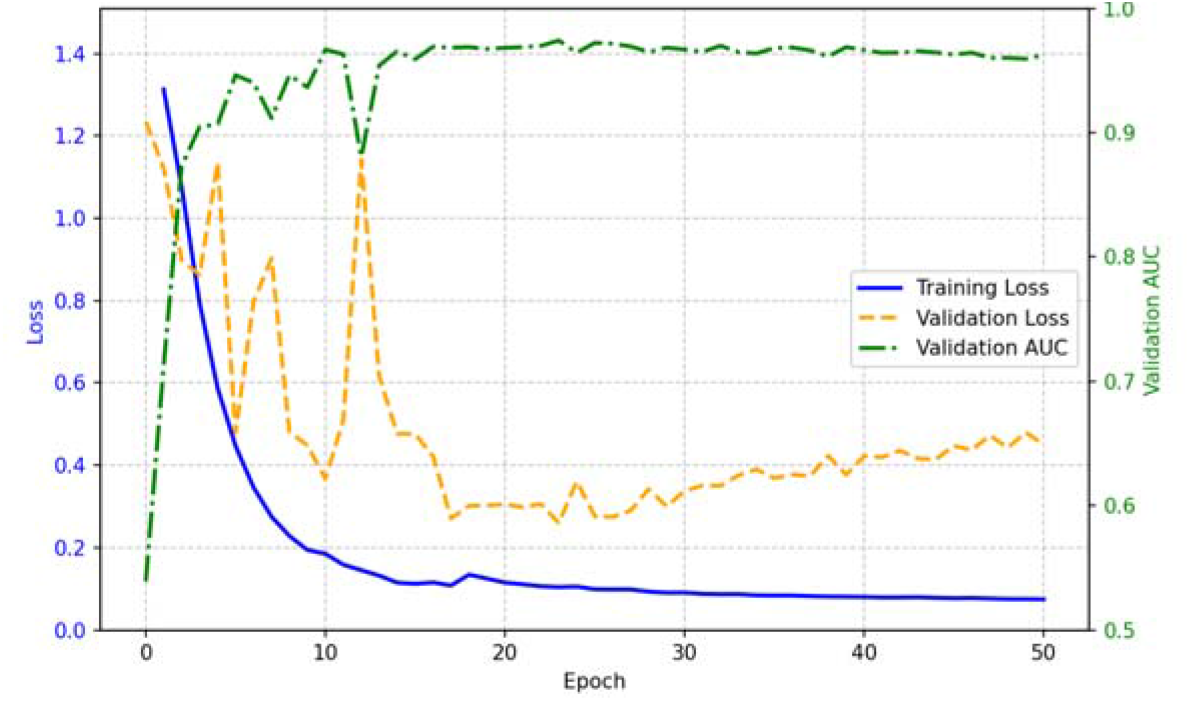
Training and validation loss, along with validation macro-AUC, across training epochs.

### Post-training Threshold Tuning

For the selected best-AUC checkpoint, we applied the same threshold tuning procedure used for the baseline model. Thresholds were optimized on the validation set using the identical grid (0.01–0.99, step size 0.05), evaluating both a single global threshold shared across all 115 labels and label-specific thresholds optimized independently. The resulting validation-set performance under both strategies is summarized in Table 2A.

**Table 2A.** Validation-set performance comparison under global versus per-label threshold tuning for the fine-tuned model (best-AUC checkpoint).

| Threshold Strategy | Metric | Macro | Macro(95%CI) | Micro | Micro(95%CI) |
| --- | --- | --- | --- | --- | --- |
| Global threshold | F1 | 0.709 | 0.694-0.718 | 0.685 | 0.675-0.697 |
|  | Sensitivity | 0.707 | 0.692-0.723 | 0.639 | 0.625-0.653 |
|  | Specificity | 0.998 | 0.998-0.998 | 0.998 | 0.998-0.998 |
|  | Precision | 0.783 | 0.773-0.793 | 0.739 | 0.727-0.751 |
| Per-label thresholds | F1 | 0.796 | 0.782-0.805 | 0.802 | 0.788-0.814 |
|  | Sensitivity | 0.801 | 0.784-0.817 | 0.815 | 0.794-0.834 |
|  | Specificity | 0.998 | 0.998-0.998 | 0.998 | 0.998-0.998 |
|  | Precision | 0.831 | 0.821-0.840 | 0.789 | 0.780-0.797 |

The thresholds determined on the validation set were subsequently fixed and applied to the test set without further tuning. The resulting test set performance is reported in Table 2B.

**Table 2B.** Test-set performance using validation-selected global and per-label thresholds for the fine-tuned model (best-AUC checkpoint).

| Threshold Strategy | Metric | Macro | Macro(95%CI) | Micro | Micro(95%CI) |
| --- | --- | --- | --- | --- | --- |
| Global threshold | F1 | 0.713 | 0.685-0.731 | 0.698 | 0.672-0.722 |
|  | Sensitivity | 0.700 | 0.679-0.721 | 0.651 | 0.626-0.676 |
|  | Specificity | 0.998 | 0.998-0.999 | 0.998 | 0.998-0.999 |
|  | Precision | 0.761 | 0.727-0.796 | 0.752 | 0.712-0.792 |
| Per-label thresholds | F1 | 0.779 | 0.756-0.794 | 0.756 | 0.737-0.773 |
|  | Sensitivity | 0.819 | 0.799-0.837 | 0.792 | 0.768-0.815 |
|  | Specificity | 0.998 | 0.997-0.998 | 0.998 | 0.997-0.998 |
|  | Precision | 0.766 | 0.741-0.792 | 0.722 | 0.694-0.752 |

To evaluate threshold calibration, we compared global and per-label strategies on the validation set (Table 2A). Based on the results, the per-label thresholding strategy outperformed the single global threshold method, with macro-F1 increasing from 0.709 to 0.796 and micro-F1 from 0.685 to 0.802. The improvement was primarily driven by higher recall, while specificity remained approximately 0.998 under both strategies. The validation-selected thresholds were then fixed and applied to the test set (Table 2B), where the same pattern was observed: macro-F1 increased from 0.713 to 0.779 and micro-F1 from 0.698 to 0.756. Given these consistent improvements across both validation and test sets, we report the per-label threshold results as the final model performance.

Because HCC coding involves severe class imbalance, with the majority of label–note pairs being negative, high specificity is expected in multi-label evaluation. Importantly, the strong recall and macro-F1 scores demonstrate that the model is not merely defaulting to negative predictions but instead achieves balanced discrimination across both common and low-prevalence HCC categories.

### Fine-Tune Performance Gain

The fine-tuned model ultimately shows significant improvement in classification performance over the non-trained model. This is based on the results, where the primary metric, macro-F1, increased from 0.017 (95% CI: 0.015– 0.019) in the untrained baseline to 0.779 (95% CI: 0.756–0.794) in the fine-tuned model, while micro-F1 increased from 0.018 (95% CI: 0.017–0.019) to 0.756 (95% CI: 0.737–0.773). In addition, per-label thresholds outperformed a single global threshold for the fine-tuned model, which shows that label-specific thresholds better accommodate prevalence heterogeneity and class imbalance across HCC codes.

### Comparison with GPT-4o (Zero-Shot and Five-Shot Settings)

We evaluated GPT-4o under both zero-shot and five-shot prompting configurations using the same test split and evaluation metrics as the fine-tuned PubMedBERT model. GPT-4o was accessed via the OpenAI API platform to ensure controlled and reproducible inference. To ensure reproducibility and output stability, each HER note was processed independently using a complete prompt per request. In preliminary experiments, we observed that batching multiple HER notes within a single API request occasionally resulted in unstable outputs, including generation of non-permitted HCC codes, omission of required codes, and formatting inconsistencies across consecutive predictions. To mitigate these issues and ensure consistent evaluation, each API call was restricted to a single target HER note.

Each prompt consisted of the following components:

1. Role specification defining the model as an HCC coding specialist under CMS guidelines
2. Task instruction requiring assignment of valid HCC codes based solely on the provided clinical note;
3. Full list of eligible HCC codes with definitions;
4. Five-shot setting continues HER–code pairs (two single-code, two multi-code, and one no-code case)
5. Target HER note;
6. Explicit output format specification

The complete prompt templates for zero-shot and five-shot configurations are presented in Appendix A.

All the performance metrics, such as macro F1, micro F1, sensitivity, and specificity, along with their respective 95% confidence intervals, have been calculated by using 500 bootstrap resamples for consistent comparisons between the models. The results for zero-shot and five-shot configurations are presented in Table 3.

**Table 3.**
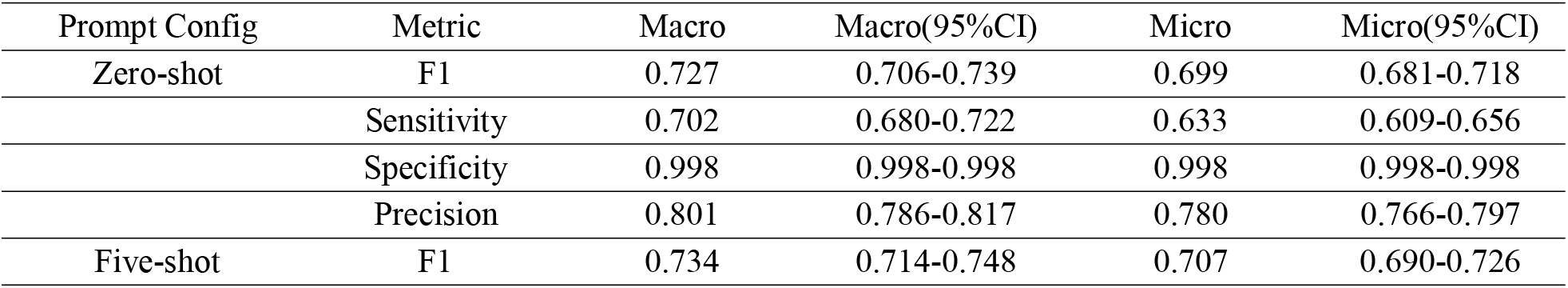
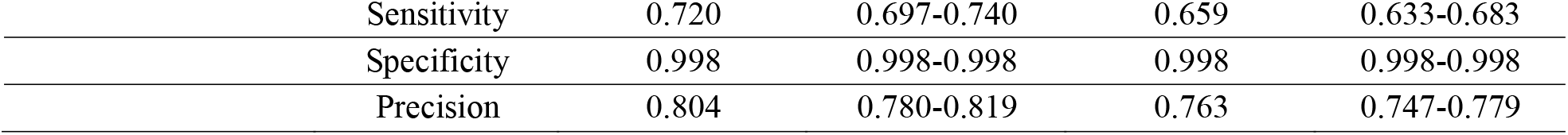
Test-set performance of GPT-4o under zero-shot and five-shot prompting.

As can be seen from the results presented in Table 3, GPT-4o performed well on the test set with both zero-shot and five-shot prompting configurations. For the zero-shot case, macro-F1 was 0.728, with micro-F1 scoring 0.699, and specificity close to 0.999. When five-shot samples were added, there was a small improvement in performance, with macro-F1 scoring 0.735 and micro-F1 scoring 0.708, mainly through a boost in sensitivity. However, the fine-tuned model had a macro-F1 score of 0.779 and a micro-F1 score of 0.756, which is approximately 4-5 percent higher than that of the five-shot GPT-4o model, with a similarly high level of specificity. Overall, fine-tuning provided more consistent performance for this imbalanced multi-label HCC classification task.

### Evaluation Platform

All experiments were performed on the Lambda Labs GPU cluster, utilizing a single NVIDIA A100-SXM4 (40GB VRAM). The test machine had 30 vCPUs, 216GB RAM, and 512GB local storage, with NFS storage available for data access. The test machine ran Ubuntu 22.04 with a kernel version of 6.8.0-1040-nvidia. The experiments were performed using PyTorch 2.1 with CUDA 12.4 and cuDNN 8.1, along with the Hugging Face Transformers library, which is a memory-efficient transformer implementation. All experiments were performed within a Docker container. A brief overview of the hardware and software environment is listed in Table 4.

**Table 4.** Evaluation platform configuration.

|  |  |
| --- | --- |
| Cluster | Lambda Labs |
| GPU | NVIDIA A100-SXM4 |
| VRAM | 40 GB |
| CPU | 30 vCPUs |
| RAM | 216 GB RAM |
| Storage | 512 GB + 8.0E external |
| OS | Ubuntu 22.04 |
| Kernel | 6.8.0-1040-nvidia |
| Frameworks | PyTorch 2.1; CUDA 12.4; cuDNN 8.1 |
| Tooling | NeMo; Transformers |
| Container | Docker |

## Discussion

### Principal Findings

Our work shows that task-specific fine-tuning significantly improves performance for HCC coding from unstructured EHR texts. Specifically, compared with the baseline where no training was performed (i.e., macro F1 ≈ 0.017), our fine-tuned PubMedBERT model achieved 0.779 in macro F1 and 0.756 in micro F1 on the test set. Moreover, our experiments suggest the importance of label-wise threshold calibration in the imbalanced multi-label setup. Overall, the performance of label-wise thresholding surpassed the baseline where a single global threshold was used, mainly because of the significant gains in sensitivity while maintaining relatively consistent specificity at around 0.998. Although GPT-4o performed well in zero-shot and five-shot settings, with macro F1 scores of 0.728 and 0.735, respectively, our fine-tuned model performed better in both macro F1 and balance, with 0.779 in macro F1 and significant gains in recall.

### Implications for Specialized Clinical Coding

The present findings underscore the distinct challenges of hierarchical condition category coding as a specialized clinical NLP task. Unlike open-ended language understanding problems, HCC classification involves highly imbalanced, multi-label prediction across numerous low-prevalence categories, with direct implications for reimbursement and risk adjustment. In such settings, recall and calibration are particularly critical, as false negatives may lead to systematic underestimation of patient complexity.

Our results highlight the importance of label-specific decision boundaries in this context. Because prevalence varies substantially across HCC categories, a single global threshold is unlikely to yield optimal trade-offs between precision and sensitivity. The smooth improvements observed with per-label threshold calibration imply that tailored decision strategies play an important role in achieving balance and reliability in coding regulation scenarios. More generally, the findings imply that domain-specific models retain significant advantages in structured and guideline-based clinical tasks. Although general-purpose language models of large capacity possess significant semantic capacity, coding scenarios may require explicit alignment with domain-specific data and decision strategies to ensure reliable and auditable outputs.

### Computational Cost and Deployment Considerations

Beyond predictive performance, important differences arise from model architecture and deployment characteristics. The fine-tuned PubMedBERT model was implemented as a direct multi-label classification system with a dedicated output layer for HCC prediction. Once trained, the model accepts raw clinical notes as input and directly outputs structured HCC codes, without requiring repeated inclusion of task instructions, coding guidelines, or reference lists at inference time.

In contrast, GPT-4o operates as a generative model and requires explicit prompt engineering to constrain behavior and enforce structured output formatting. In our experiments, each API call included the full HCC reference list and coding rules, and in the five-shot configuration, additional demonstration examples. Across all requests (zero- and five-shot combined), a total of 5.1 million input tokens were consumed, corresponding to an empirical average of approximately 1,800 input tokens per note and a cost of roughly $0.0027 per instance. While modest at a small scale (approximately $9.7 for 3,600 notes), inference cost scales linearly with dataset size and increases further under few-shot prompting due to additional demonstration tokens.

Fine-tuning required approximately 1.5 hours of GPU time on a $1.29/hour instance (total training cost ≈ $1.94). After training, inference over 3,600 test notes required only 9.4 seconds (≈383 notes per second), resulting in negligible marginal cost per note. Importantly, once trained, the classification model can be deployed locally, including on standard hardware without dedicated GPU resources which allows near-zero per-instance inference cost and high-throughput batch processing.

Operational considerations further differentiate the approaches. API-based inference may be subject to account-level rate limits and request caps; in our case, a new account was restricted to one request every 20 seconds during initial usage, which introduced additional time constraints independent of model computation. Locally deployed models, by contrast, provide full control over throughput, versioning, and system configuration, which may simplify integration into institutional clinical workflows and large-scale retrospective coding applications.Collectively, these findings suggest that while large generative models demonstrate strong zero- and few-shot performance, domain-adapted classification models offer advantages in cost predictability, scalability, and deployment control for high-volume, structured clinical risk-adjustment tasks such as HCC coding.

## Future Directions

Future work will proceed along three complementary directions. First, we aim to expand the labeled corpus beyond the current 36,000 ICU notes to improve statistical power and strengthen external validity across institutions and patient populations. Larger-scale annotation will enable more robust estimation of rare HCC categories and further reduce class imbalance effects.

Second, we plan to explore fine-tuning of larger, open-weight language models (e.g., Qwen-7B) to better characterize the performance–efficiency trade-offs between medium-scale domain-adapted transformers and more expressive generative architectures. This will help delineate the practical performance boundary of instruction-tuned LLMs under structured clinical coding constraints.

Third, we aim to deploy the model in practical healthcare environments to test its influence on the efficiency of the coding workflow, the precision of risk adjustment, and the level of human oversight needed by clinicians. Future testing in practical environments will allow for an assessment of the model’s stability, calibration, and the human-AI interaction process.

## Conclusion

We present HCC-Coder, a transformer-based framework for automated hierarchical condition category (HCC) classification from unstructured EHR narratives. By fine-tuning PubMedBERT with an imbalance-aware inverse-frequency weighting scheme and label-wise threshold calibration, the proposed approach achieves strong performance on a MIMIC-IV–derived benchmark, attaining macro- and micro-F1 scores of 0.779 and 0.756 on the independent test set. This demonstrates that domain-adapted transformer models can effectively translate free-form clinical documentation into structured outputs that match healthcare risk adjustment workflows. Besides prediction accuracy, HCC-Coder provides a reproducible evaluation benchmark for multi-label clinical coding, a scalable human-LLM hybrid annotation pipeline, and a rigorous framework for label-dependent decision calibration in imbalanced scenarios. The combination of these elements provides a useful foundation for scalable and trustworthy clinical coding automation. The results again stress the need for domain adaptation and task-oriented optimization of language models, especially when used in structured, regulation-sensitive healthcare applications.

## Data Availability

All data produced in the present work are contained in the manuscript
All data produced are available online at

